# Creation and evaluation of full-text literature-derived, feature-weighted disease models of genetically determined developmental disorders

**DOI:** 10.1101/2021.11.04.21265878

**Authors:** T.M Yates, A. Lain, J. Campbell, T.I. Simpson, D.R. FitzPatrick

## Abstract

There are >2500 different genetically-determined developmental disorders (DD), which, as a group, show very high levels of both locus and allelic heterogeneity. This has led to the wide-spread use of evidence-based filtering of genome-wide sequence data as a diagnostic tool in DD. Determining whether the association of a filtered variant at a specific locus is a plausible explanation of the phenotype in the proband is crucial and commonly requires extensive manual literature review by both clinical scientists and clinicians. Access to a database of weighted clinical features extracted from rigorously curated literature would increase the efficiency of this process and facilitate the development of robust phenotypic similarity metrics. However, given the large and rapidly increasing volume of published information, conventional biocuration approaches are becoming impractical.

Here, we present a scalable, automated method for extraction of categorical phenotypic descriptors from full-text literature. Papers identified through literature review were downloaded and parsed using the Cadmus custom retrieval package. Human Phenotype Ontology terms were extracted using MetaMap, with 76-83% precision and 72-81% recall. Mean terms per paper increased from 9 in title + abstract, to 69 using full text. We demonstrate that these literature-derived disease models plausibly reflect true disease expressivity more accurately than gold standard manually-curated models, through comparison with prospectively gathered data from the Deciphering Developmental Disorders study. AUC for ROC curves increased by 5-10% through use of literature-derived models. This work shows that scalable automated literature curation increases performance and adds weight to the need for this strategy to be integrated into informatic variant analysis pipelines.

## Introduction

The use of genome-wide sequencing technologies combined with rational frequency, inheritance and consequence-based variant filtering strategies has transformed the diagnosis of genetically-determined developmental disorders (GDD) (1–4). Although filtering efficiently reduces the number of genetic variants for diagnostic consideration, each of these have to be reviewed to determine if the clinical features (phenotype) of the individual being tested can be explained by one or more of these genotypes. This usually requires manual review of peer-reviewed case reports/series which describe relevant genotype/phenotype associations (5). The matching is based on expert opinion as few metrics exist to rank the associations with any statistical rigor.

Phenotypic data in the literature is not usually recorded in a standardised, computationally-tractable format. Plain text descriptions of similar clinical features may be recorded in several different ways. For example, a technical term such as ‘hypertelorism’, may be recorded as its synonym ‘widely spaced eyes’. In addition, case reports are found across a wide range of journals, with different structures and file formats for each publication.

The Human Phenotype Ontology (HPO) was developed to store phenotypic data in a computationally-accessible format (6). Several initiatives have been developed to link diseases to phenotype data, in the form of HPO terms, for example OMIM (Online Mendelian Inheritance in Man) (7) and Orphanet (8). However, these rely on manual expert curation and therefore are not inherently scalable, and cannot be updated automatically.

Methods of extracting phenotype data from text at scale previously have relied on abstracts or open access papers (9,10). At the time of writing, Europe PubMed Central (EPMC, https://europepmc.org/) contained approximately 39.5 million articles, of which only 3.8 million were open access. Therefore, there is likely a significant volume of phenotypic data which has not been used previously.

Our overall aim is to create systems that allow scalable, automated and clinically orientated literature curation to aid the robustness of diagnosis through genomic testing. Here, we present a method for creating disease models describing GDD, comprising lists of HPO terms, with weighting according to term frequency in the literature. Utilising intellectual property law for research in the UK (11), we retrieve the full text of almost all relevant case reports, and extract phenotypic data mapped to HPO terms for a set of GDD. We evaluate this against prospectively gathered patient phenotypes from the Deciphering Developmental Disorders (DDD) study (1), and compare to current gold standard manual curation.

## Materials and methods

### Full text retrieval

A test set of 99 GDD defined in the Developmental Disorders Genotype2Phenotype (DDG2P) database (4) were selected, to include conditions well-represented in the DDD study (1) and in OMIM (7). For each of these, a literature review was undertaken using PubMed searches. The initial search was by gene symbol in title - {gene symbol}[TI]. If this returned less than 300 results, an abstract review was undertaken to identify relevant case reports. If the initial search returned more than 300 results, modifier terms were added such as {gene symbol}[TI] AND {syndrome name} or {gene symbol}[TI] AND ‘intellectual disability’. Only papers which described case reports for variants in a single gene were included.

From this process, a list of case report PubMed IDs (PMIDs) was generated for each of the 99 diseases. These were inputted into the Cadmus full text retrieval package (https://doi.org/10.5281/zenodo.5618052), which will be described in detail in a forthcoming publication. In brief, metadata obtained using each PMID was used to send requests for download for each paper to sources which authorise full text retrieval for research purposes. Multiple sources were used to maximise the chances of download, including, but not limited to, Crossref, doi.org and Europe PubMed Central (EPMC). File formats generated through this include HTML, XML, PDF and plain text. Where multiple formats were retrieved, a series of quality assessments were used to identify the best full text version. The text was cleaned and converted to a string. The abstract and references were parsed out. This final document was used for all following steps and will be defined as the ‘full text’ in subsequent paragraphs.

### Phenotype extraction

Phenotypic features in text were identified and mapped to Unified Medical Language System (UMLS®) concept unique identifiers (CUIs) using the 2018 release of MetaMap (12). The source vocabulary for MetaMap was restricted to CUIs corresponding to the Human Phenotype Ontology (HPO) (6), which is Category 0 under the UMLS Metathesaurus® licence. MetaMap includes negation information for each phenotypic feature identified; the frequency of each non-negated feature in the text was used for term weighting. Negated terms, whilst potentially useful to identify phenotypic features which are not associated with a given disorder, were not included in this work as the comparison DDD dataset does not include these. CUIs were then mapped to HPO terms, using the mappings in the UMLS Metathesaurus® (Release 2020AA).

### MetaMap performance evaluation

To test the performance of MetaMap (12), a set of 50 papers – randomly selected from the list of full text downloads described above – were manually annotated. Non-negated phenotypic features in the text were annotated directly to HPO terms. Against this standard, MetaMap was evaluated for precision (fraction of true positive terms in output) and recall (fraction of true positive terms compared to all true terms in full text), using a variety of usage options. The F1 score (harmonic mean of precision and recall) was also calculated.

### Disease model creation

For each group of PMIDs corresponding to a given disease, extracted HPO terms and their frequency were aggregated to create a ranked, weighted disease model. Corresponding diseases in manually curated sets were mapped using the disease MIM identifier, and the HPO annotated file genes_to_phenotype file downloaded from http://purl.obolibrary.org/obo/hp/hpoa/genes_to_phenotype.txt on the 22^nd^ of April 2021. This includes disease-specific HPO term lists (models) from OMIM (mim2gene) (7) and Orphanet (8). OMIM-derived terms mostly do not include frequency/weighting, whereas Orphanet terms are uniformly annotated. The frequency in both cases was recorded as an HPO frequency term e.g. Very frequent (HP:0040281); present in 80 to 99% of the cases. OMIM models corresponding to the 99 G2P/DDD disease set were used for the majority of analyses. A subset of 43 Orphanet models were used for weighted analyses, as the Orphanet annotations were not available for the full set. HPO term models from the DDD study (1) were created using aggregated lists from probands with diagnoses corresponding to the 99 disease set. These were recorded prospectively by clinicians recruiting individuals to the study with a suspected, undiagnosed GDD.

### Disease model evaluation

Evaluation of literature-derived models through comparison with DDD models was designed to assess similarity to real life, clinical, prospective data. OMIM models were used as an example of gold standard manual curation (7). Two similarity metrics were used: rank biased overlap (13) and semantic similarity using information content (IC) as defined by Resnik (14).

Rank biased overlap (RBO) is a method of comparing ranked lists which is top-weighted, can compare lists which contain differing members, and is monotonic with increasing depth of list (13). RBO allows for the top-weightedness parameter p to be fine-tuned to weight the score more or less towards higher-ranked items in the list. For this analysis, p was equal to 0.98, weighting towards the top 50 terms in a list. RBO may be expressed as a min-max range, however for this work the extrapolated RBO_EXT_ point score was used for ease of comparison. RBO was calculated for all models in the literature-derived set vs the DDD set, OMIM vs DDD and literature vs OMIM (1,7). Literature- and DDD-derived models were ranked according to term frequency. OMIM model terms are not generally annotated with a frequency; the frequency of each term across all OMIM models was used for ranking.

For the semantic similarity measure, the HPO terms in both comparison datasets, e.g. literature vs DDD were used to calculate the IC for each term following the method used by Helbig et al. (15). If f is the number of diseases annotated with an HPO term g, and n is the total number of diseases, the IC_g_ is defined as – log_2_(f/n) (16). The most informative common ancestor (MICA) of two terms is the parent term in the ontology with the highest IC. For a disease-disease comparison, a matrix *m* is created with HPO terms of one disease (*l* terms) as the rows, and the terms of the other (*k* terms) as the columns. Each position in the matrix (*m*_*ij*_) is a comparison between pairs of HPO terms, and is populated with the MICA for that pair. The similarity score between diseases is computed by summing the average of the rows and the columns, with a normalisation measure (15).

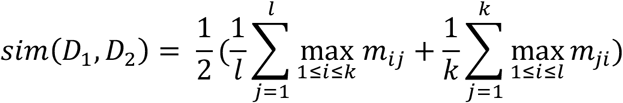

Semantic similarity scores between literature-derived, DDD and OMIM sets were performed using unweighted models, i.e. every term appeared uniquely per model.

A comparison of weighted models using the Orphanet (8) subset was undertaken, however there was no straightforward method of normalizing the frequency models between datasets. For the Orphanet models, which consisted of a flat list of phenotype terms annotated with HPO frequency terms, the percentage range in HPO frequency annotation was mapped to the mean of this range. For example, a term annotated as Very frequent with the range 80 to 99% was mapped to 89. Each term in a disease list was repeated according to its frequency mapping, thereby creating a weighted model. For literature-derived and DDD models (1), frequency annotations were binned into four bins using numpy.histogram, corresponding to HPO terms Very frequent, Frequent, Occasional and Very rare. Frequency weighting was then applied as per the Orphanet models. For models weighted in this manner, *l* row terms and *k* column terms in disease comparison matrix *m* therefore may contain repeats, with the MICA sum average altering accordingly (17). Comparisons were calculated for the 43 diseases in the Orphanet set vs DDD models, and for the corresponding 43 literature-derived models vs DDD.

## Results

Full text papers describing GDD were downloaded, phenotypic features extracted and weighted disease models constructed. These were evaluated against data from the DDD study (1) and gold standard manually curated models. An overview of this process is shown in Figure 1.

**Figure 1.**
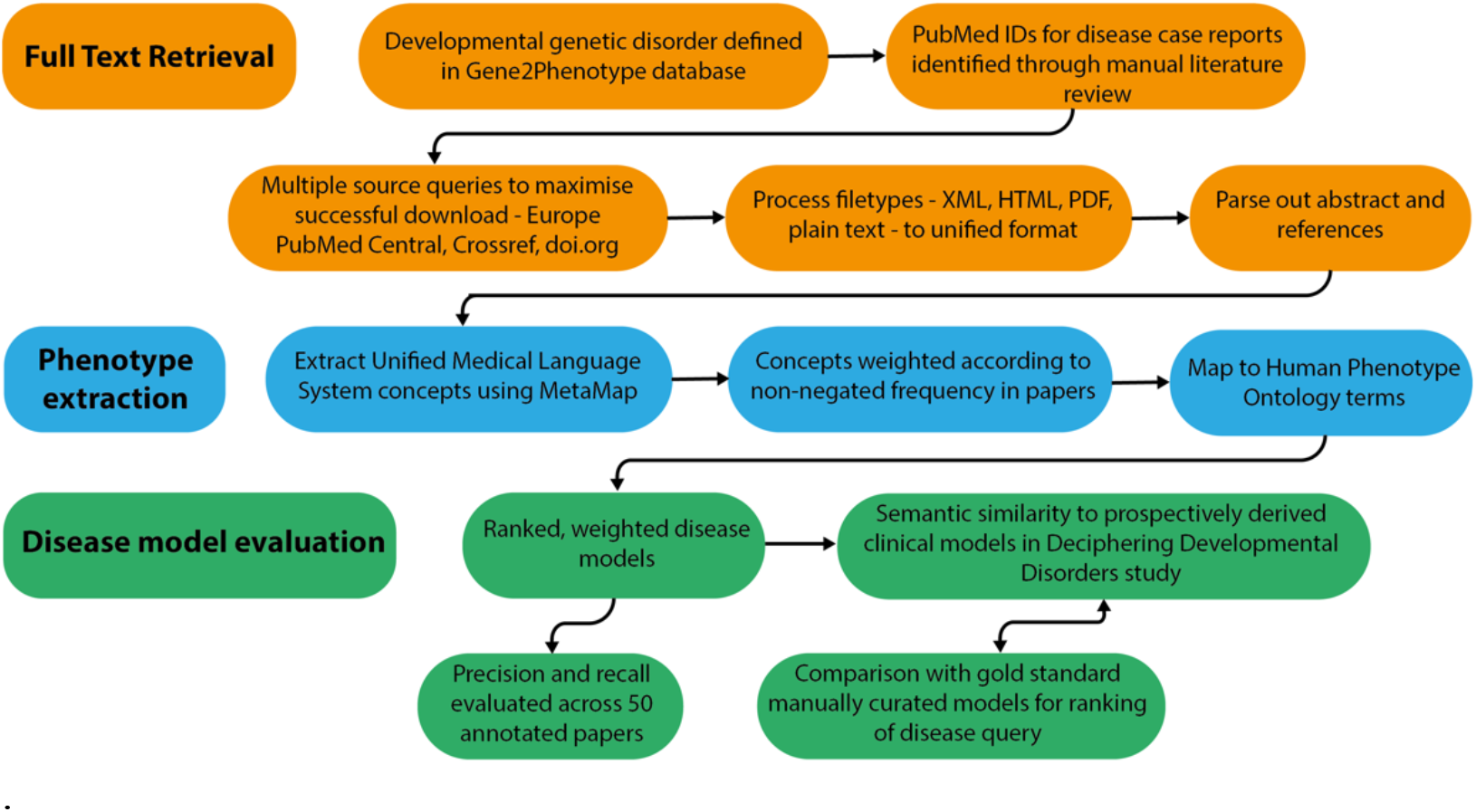
Overview of disease model pipeline. Input was PubMed IDs for case reports describing developmental genetic disorders. Full text downloads were performed using the Cadmus package. Output was disease models consisting of Human Phenotype Ontology terms weighted according to their frequency in full text. These were evaluated against ‘real life’ models from the Deciphering Developmental Disorders study, and against gold standard manually curated models

### Full text retrieval and phenotype extraction

For 99 GDD in the test set, 1018 relevant case reports were identified. Cadmus (https://doi.org/10.5281/zenodo.5618052) successfully downloaded at least one format (HTML/XML/PDF/plain text) for 962/1018 papers (94.5%) (Supp. Table 1). There were significantly more HPO terms in full text than in title + abstract after phenotype extraction (Supp. Table 2). There were also more terms in the full text-derived models than in OMIM (Figure 2). Table 1 shows the number of unique terms across the whole dataset by source.

**Figure 2.**
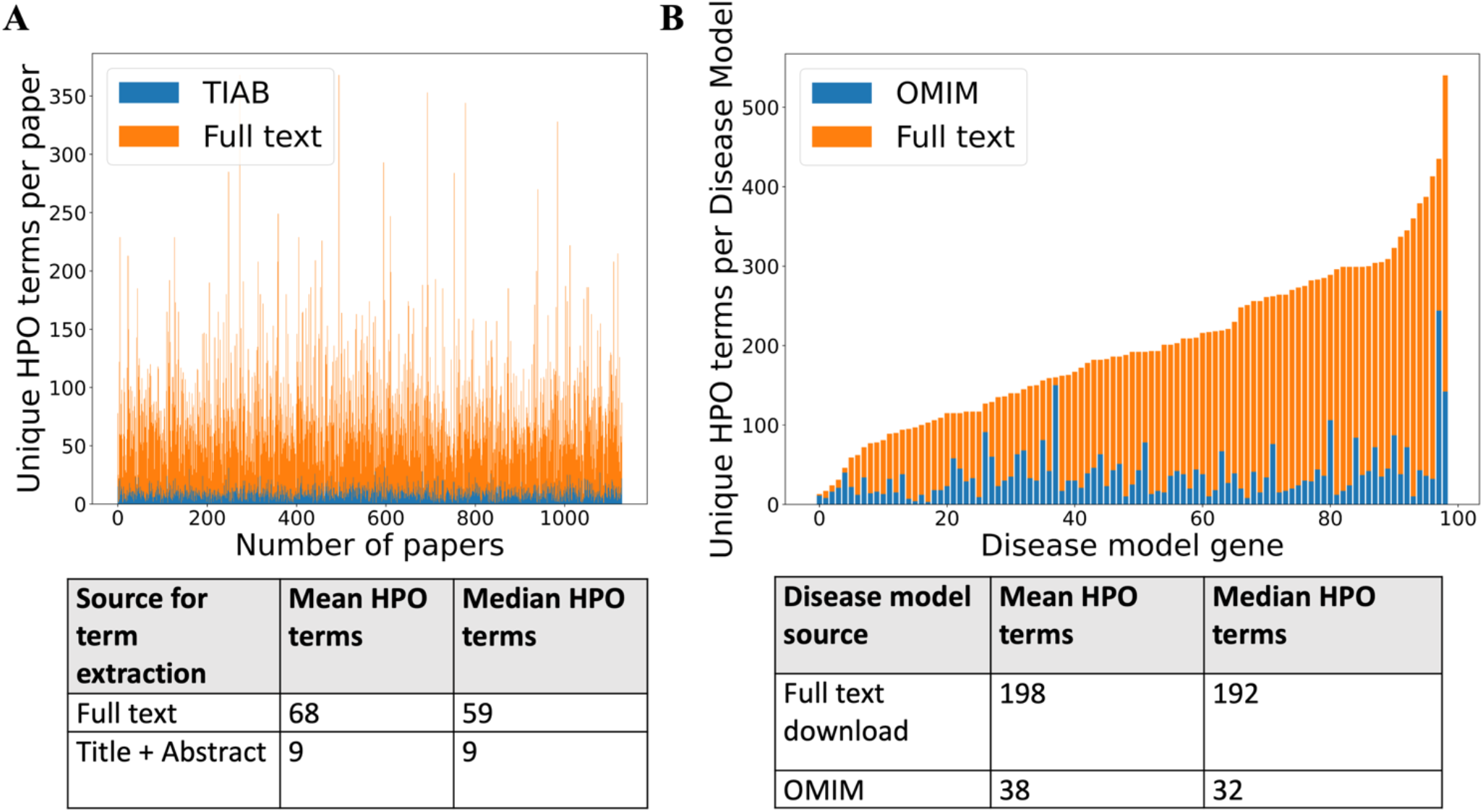
(A) Comparison of number of unique HPO terms extracted from full text vs title + abstract for sample of 962 papers, using full text download pipeline. (B) Comparison of number of unique HPO terms in disease models generated by full text download pipeline vs manually curated models in OMIM, for sample of 99 diseases.

**Table 1.** Number of unique HPO terms across 99 disease test set

| Source | Full text download | DDD | OMIM | Full text + DDD | DDD + OMIM | Full text + DDD + OMIM |
| --- | --- | --- | --- | --- | --- | --- |
| Number of unique HPO terms in set | 3234 | 1569 | 972 | 3741 | 1935 | 3864 |

The performance of phenotype concept extraction and HPO mapping in a 50 paper sample from the above set was evaluated using precision, recall and F1 score for a number of different MetaMap (12) output options (Table 2). Restricting source vocabulary to HPO alone was chosen as the performance was similar to other options, but with a quicker processing speed. This configuration was used for all the other analyses in this work.

**Table 2.** Performance of MetaMap usage options. Evaluated against 50 manually-annotated papers describing developmental disorders. Word sense disambiguation – disambiguate concepts with similar scores. Restrict to HPO – use only human phenotype ontology for mapping concepts. No derivational variants – compute word variants without using derivational variants. Blanklines off – process text as whole document. Conjunction processing – join conjunction-separated phrases.

| MetaMap options | Precision | Recall | F1 score |
| --- | --- | --- | --- |
| Word sense disambiguation, no derivational variants, restrict to HPO | 0.795 | 0.785 | 0.790 |
| Word sense disambiguation, restrict to HPO | 0.805 | 0.772 | 0.788 |
| No derivational variants, restrict to HPO | 0.789 | 0.781 | 0.785 |
| Blanklines off, restrict to HPO | 0.758 | 0.811 | 0.784 |
| Restrict to HPO | 0.775 | 0.791 | 0.783 |
| Blanklines off, word sense disambiguation, no derivational variants, conjunction processing, restrict to HPO | 0.785 | 0.776 | 0.781 |
| Blanklines off | 0.820 | 0.742 | 0.779 |
| No options | 0.836 | 0.723 | 0.776 |
| Conjunction processing, restrict to HPO | 0.767 | 0.778 | 0.772 |

### Evaluation of disease models

The comparison of weighted disease models constructed from full text to models derived from other sources was not straightforward. The example model for *CHD7* in Figure 3 illustrates some of the issues. This describes the condition CHARGE syndrome, which is an acronym for Coloboma of the eye, Heart defects, Atresia of the choanae (choanal atresia), Restriction of growth and development, and Ear abnormalities/deafness. The top 5 ranked terms in the full text-derived model are therefore highly relevant to this condition. However, a number of terms clinically relevant to these were also present in the same model, and this pattern is repeated across comparison datasets. This shows that similar phenotypic features may be recorded in a heterogenous manner, and complicates comparison between models. It is possible to use the ontology structure in HPO to relate similar terms, as per the semantic similarity MICA method. However, two terms which appear clinically similar may only share a high-level ancestor, resulting in a significant loss of informativity.

**Figure 3.**
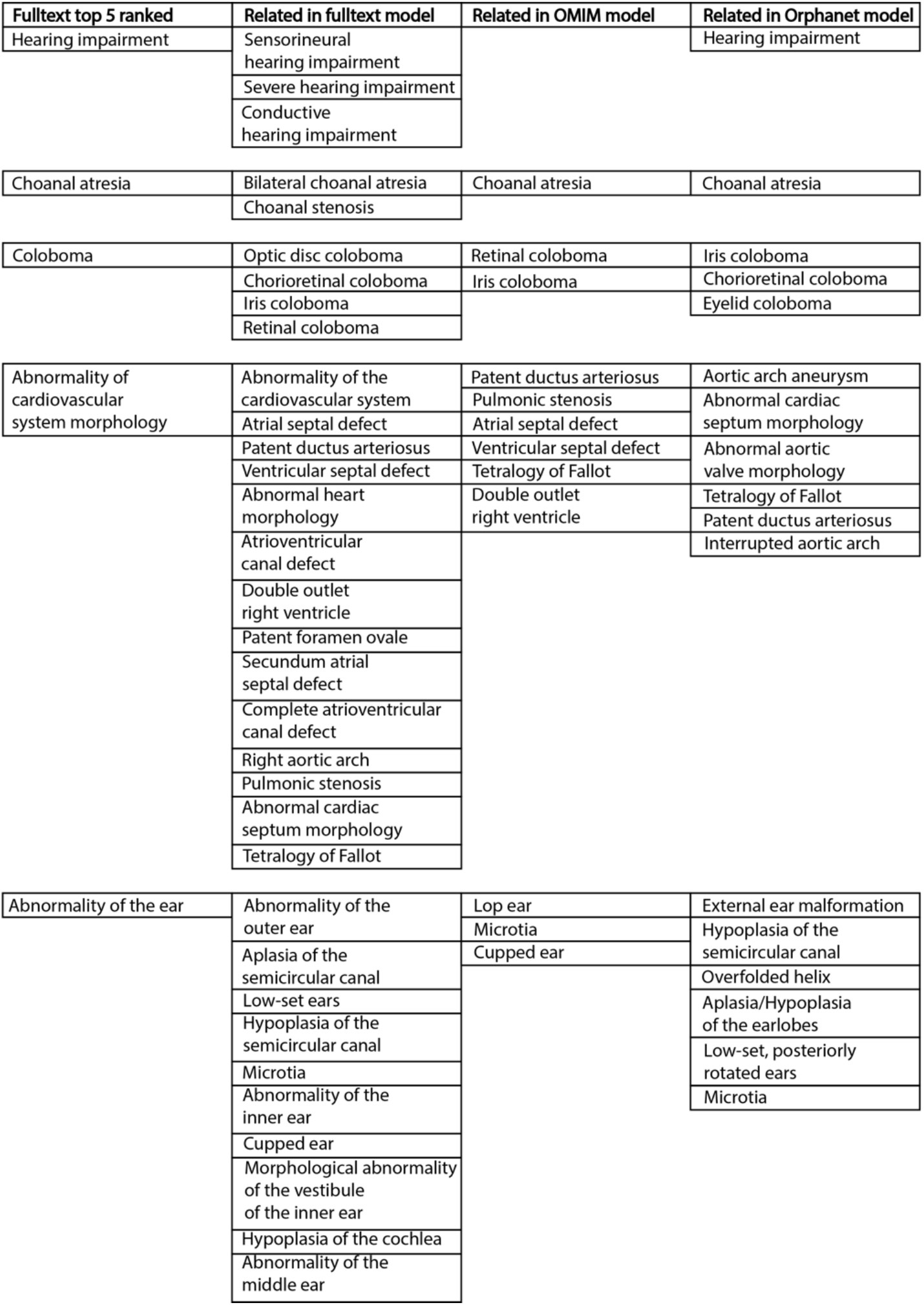
Example top five ranked terms in disease model for *CHD7/* CHARGE syndrome (left column). Clinically related terms in remainder of disease model (n=540), OMIM model (n=71) and Orphanet model (n=82) shown.

Comparison heatmaps using RBO for literature-derived, DDD and OMIM datasets (1,7) (Figure 4) were generated, where every disease in one dataset is compared to every disease in the other. This was to determine if a corresponding pair e.g. *CHD7*-full text vs *CHD7*-DDD is more similar than any other in the comparison. These show a weak, but recognizable signal for literature vs DDD and OMIM vs DDD. There is a clear signal for literature vs OMIM models, showing that full text-mined models are similar to gold standard manual curation.

**Figure 4.**
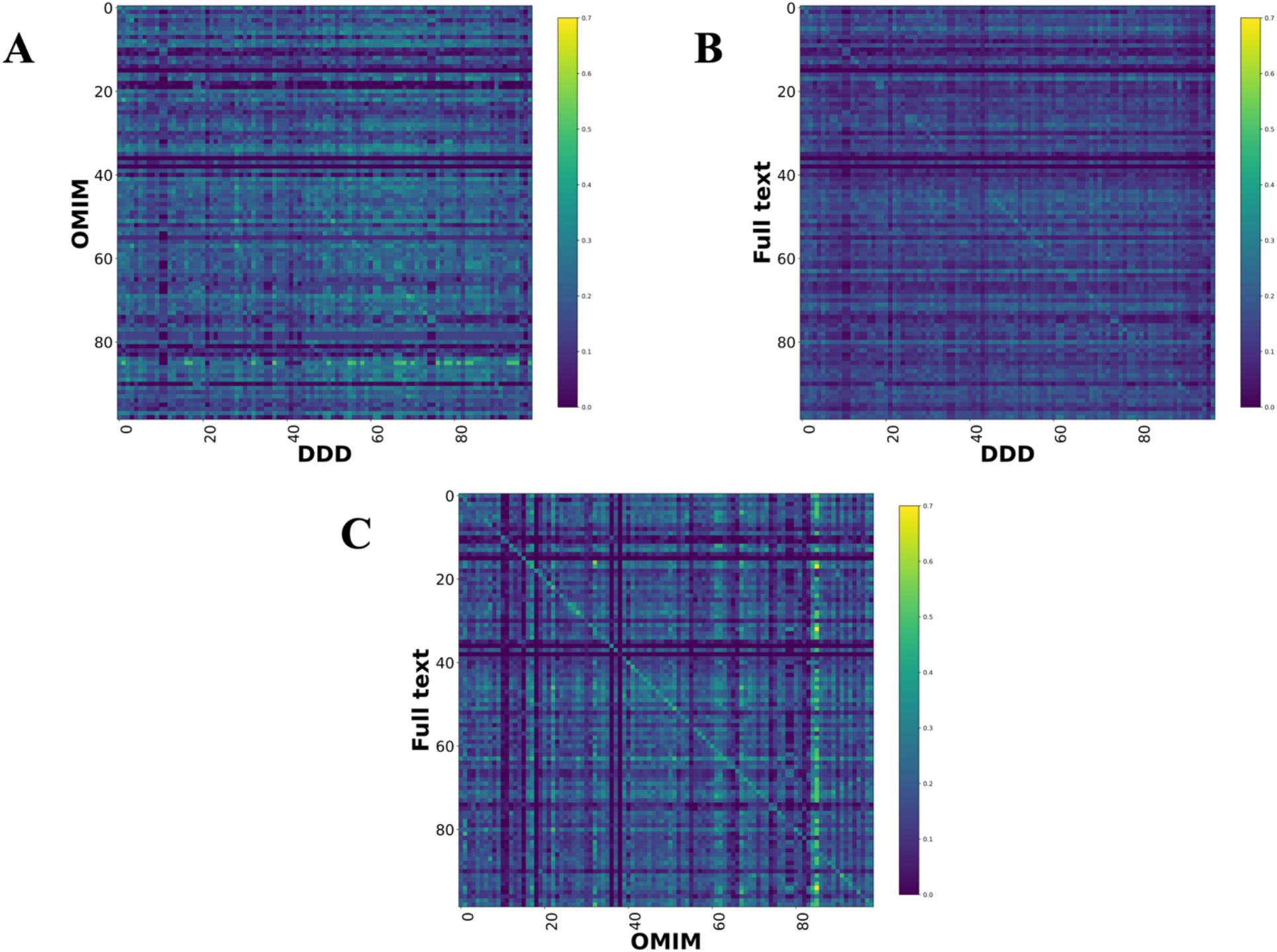
Disease model comparison heatmaps using rank biased overlap for literature-, OMIM- and DDD-derived models. Each model on the y axis is compared with every model in the DDD set. Disease/DDD models describing the same disorder are on the rightward slanting diagonal. (A) compares OMIM and DDD models. (B) compares literature-derived and DDD models. (C) compares literature-derived and OMIM models. 99 diseases in comparison set.

The performance of full text-derived models vs OMIM models in ranking the correct corresponding model in the DDD set was evaluated using ROC curves (Figure 5). The full text-derived models outperformed OMIM in both similarity metrics – ranked lists using RBO (13) and unweighted semantic similarity (15) – as defined by an increase in the area under the curve. A similar semantic similarity analysis using a subset of models from Orphanet showed comparable performance to those derived from full text when unweighted. However, weighted models did not show similar performance for either Orphanet or the full-text derived set (Supp. Figure 1.

**Figure 5.**
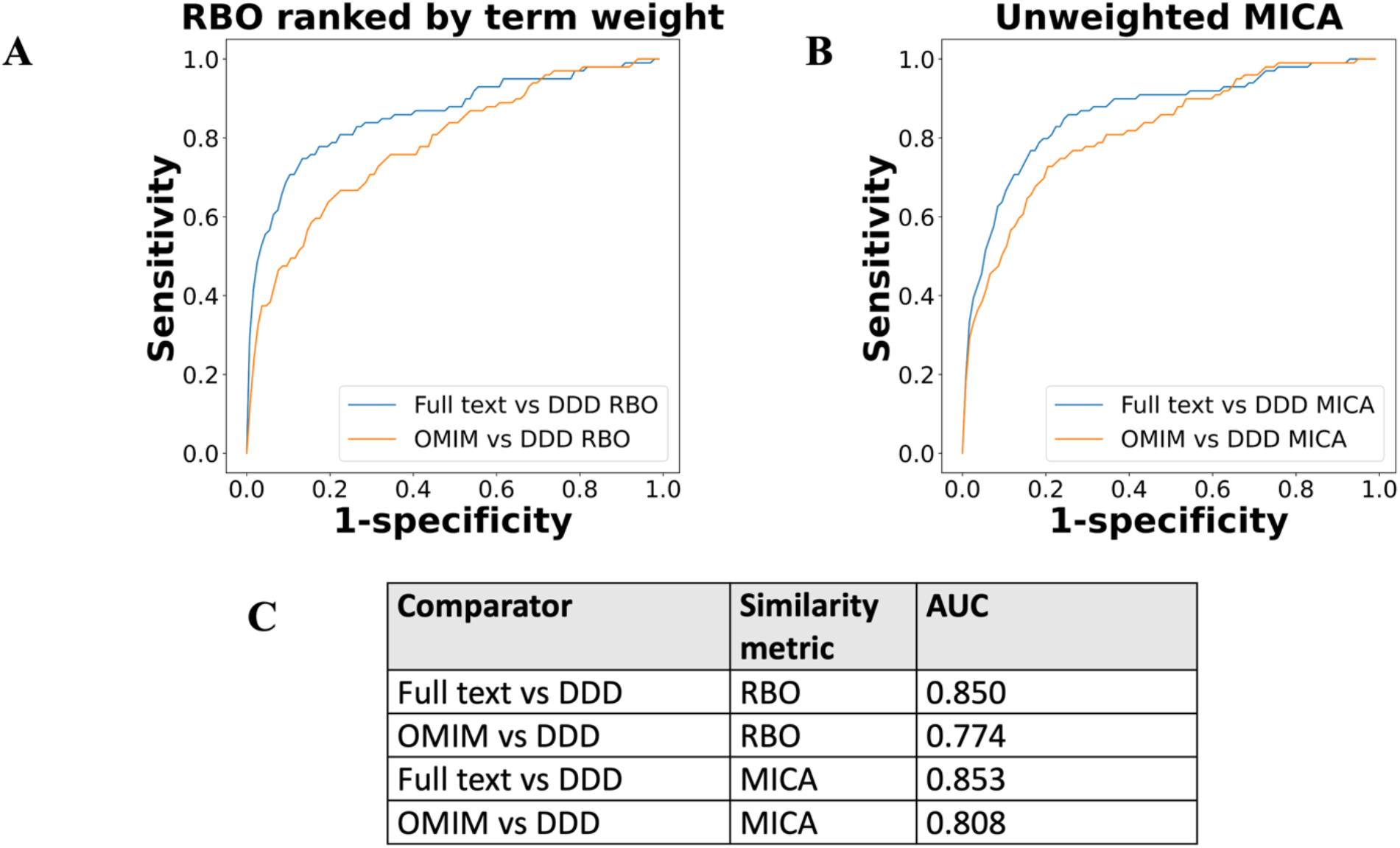
ROC curves using threshold ranking for literature-derived/OMIM disease models compared to real life terms in DDD study, across sample of 99 diseases, with a disease model and DDD model for each. Each disease model is compared to every model in the DDD set. (A) uses ranked biased overlap (RBO) to compare ranked lists of terms. Literature-derived and DDD models were ranked according to model term frequency. OMIM models were ranked according to frequency of terms across all OMIM models. (B) uses mean most informative common ancestor (MICA) to compare models, with information content calculated according to Resnik. Unweighted models were used for comparison, meaning each term in a model appeared only once, and term frequencies were not utilised. (C) shows area under curve (AUC) for each model comparison.

## Discussion

Here, we demonstrate a method for automated literature curation that enables the creation of disease models based on ranked and weighted lists of clinical features from any of the most widely used structured vocabularies. We have tested this approach on a subset of GDD to generate models using the Human Phenotype Ontology.

We showed that full text download can be achieved using standard licence agreements with both the journals and the online search engines at scale for a corpus of over 1000 papers, with close to 95% retrieval rate. Unsurprisingly, our annotation using full text returned more phenotypic descriptors per paper when compared to title + abstract, and per disease cf. manually curated models.

We showed that full text models outperform manual curation when identifying corresponding diagnoses from the DDD study, using two similarity metrics. The first of these, RBO, compared ranked lists of terms, and therefore did not utilise the ontology structure. It was perhaps surprising to show similarity between disease models without the advantages of an ontology-based metric. This may indicate that there was sufficient overlap between commonly used/prioritized terms for a given disease, despite the different methods of recording these across the datasets used, to allow for meaningful comparison. The performance improvement using full text-derived models may reflect a higher number of disease-relevant terms per disease compared to OMIM, given the disparity in model sizes between them.

The second MICA-based similarity metric used the ontology structure of the HPO to compare terms. This measure should provide a more robust method of comparison than RBO as the ontology allows for a direct measure of the relatedness of a term pair. The performance of full text-derived models was better than OMIM using this metric, even though term weighting was not utilised. However, the ROC curve derived from this was remarkably similar to the RBO based measure. It is likely that developing condensed full text-derived disease models, where clinically similar or discriminant terms are collapsed together, will demonstrate an improvement in predictive power using MICA-based similarity scoring. This task is not straightforward, as shown in Figure 3. We, and others, are currently working on methods to collapse clinically similar HPO terms without losing information.

Adding weighting, in the form of term frequency, to the MICA-based similarity comparison did not improve performance (Supp. Figure 1). This may be partly due to the challenge of normalizing term weighting between full text-derived, DDD, and manually curated datasets. The distribution of terms differed significantly between models, with generally much higher mean and median terms in the full text-derived set. Improving phenotype extraction to identify terms on a per individual, rather than per paper, basis would allow for straightforward normalizing of term weighting, enable better comparison with manually curated models, and potentially improve predictive power.

Whilst the results we present here are encouraging with regard to the performance of automated literature curation, further improvements to disease models are needed before these can replace or minimise expert clinical interpretation of the peer-reviewed literature. This includes parsing out clinical descriptive text from papers to remove superfluous phenotypic descriptors in the introduction and discussion. Upweighting of disease-discriminant terms may also be helpful, as well as collapsing clinically similar terms as mentioned above. Including negated terms, i.e. phenotypic descriptors which are explicitly never present in a disorder, may yield further improvements.

The performance of MetaMap in named entity recognition (NER) demonstrated here is comparable to newer deep learning models such as BioBert (18). This may be because MetaMap has been specifically designed – and regularly updated – to perform in the biomedical concept extraction domain. However, there could be improvements in precision and recall available through use of a deep learning NER model. Additionally, representation of phenotypic features as word embeddings could allow for novel relationships between phenotypes and disease models to be elucidated. The results presented here showing that a non-ontology based measure (RBO) is a useful similarity metric in the phenotype-disease space further support this.

Here we have demonstrated and tested a method for generating Human Phenotype Ontology-based disease models for a small subset of the >2500 different GDD. However, it is clear that this approach can be applied to any set of diseases for which there is a reasonable level of aetiological homogeneity within case reports and/or case series. We consider this approach to be easily scalable with the main bottleneck being in the efficient identification of the relevant clinical papers using online searches. We and others are actively developing systems to improve the discriminative power of the both the search strategies and the subsequent classification of the title/abstract/full text to minimise the requirement for manual review prior to full model creation. It is likely that leveraging annotated full text downloads as demonstrated here will be useful in the classification of relevant papers for phenotype extraction.

Scaling up the method shown here will enable automatic addition of new disorders, as well as updates to the phenotypic spectrum of known conditions. This means curation of databases such as DDG2P should become significantly less time- and resource-intensive. Ultimately, the expansion of automated curation to all GDD should enable improvements to existing diagnostic systems relying on manual curation (19), as well as incorporation of in-depth phenotypic data to genomic bioinformatic analysis pipelines.

## Data Availability

All data produced in the present study are available upon reasonable request to the authors

https://doi.org/10.5281/zenodo.5618052

## Acknowledgments

We thank the DDD study for access to the aggregated phenotype information on specific genetic disorders. The DDD study presents independent research commissioned by the Health Innovation Challenge Fund [grant number HICF-1009-003]. This study makes use of DECIPHER (http://www.deciphergenomics.org), which is funded by Wellcome. See Nature PMID: 25533962 or www.ddduk.org/access.html for full acknowledgement. MetaMap is provided courtesy of the U.S. National Library of Medicine.

## Supplemental Data

**Supplemental Figure 1.**
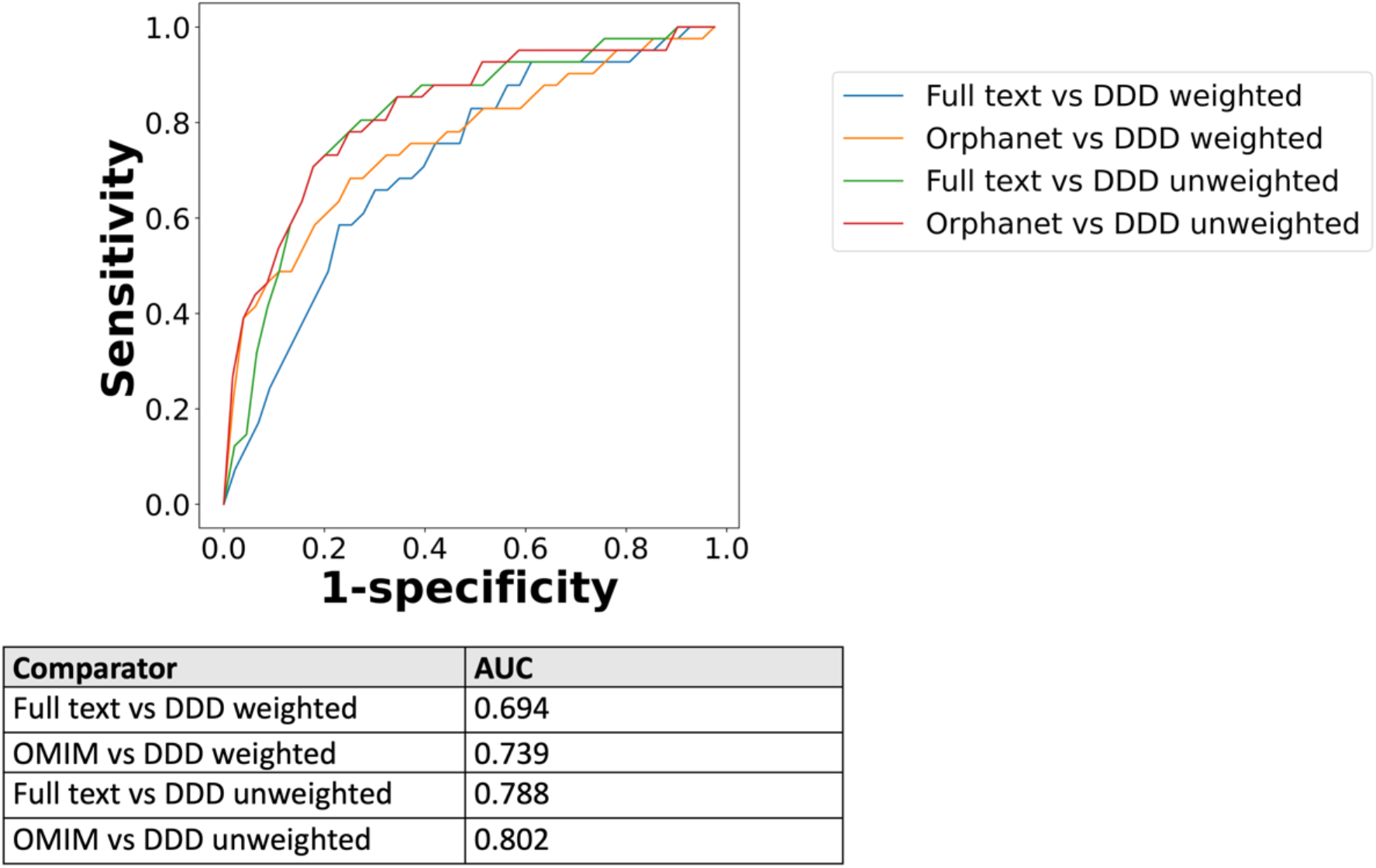
Precision curve using threshold ranking for full text-derived/Orphanet disease models compared to real life terms in DDD study, across sample of 41 diseases. Orphanet models are weighted by HPO term frequency e.g. HP:0040281 (Very Frequent 99-80%). The full text and DDD model weightings were binned according to the mean of each HPO term frequency range.

**Supplemental Table 1.** Number of full text downloads from Cadmus pipeline, for list of 1018 PMIDs corresponding to relevant case reports. There were 962 PMIDs where there was a successful download in at least one format. The total number of downloads is higher due to cases in which the same paper has been downloaded in multiple formats.

| File format | Number of full text downloads |
| --- | --- |
| PDF | 643 |
| HTML | 527 |
| XML | 233 |
| Plain text | 270 |
| Total | 1673 |

